# Factors influencing long-term recovery in critically ill COVID-19 survivors: A prospective multicentre cohort study

**DOI:** 10.1101/2024.05.01.24306267

**Authors:** Ingrid Didriksson, Attila Frigyesi, Martin Spångfors, Märta Leffler, Anton Reepalu, Anna Nilsson, Martin Annborn, Anna Lybeck, Hans Friberg, Gisela Lilja

**Affiliations:** Department of Clinical Sciences, Anaesthesiology and Intensive Care, Lund University, Sölvegatan 19, SE-223 62, Sweden; Intensive and Perioperative Care, Skåne University Hospital, Carl-Bertil Laurells gata 9, SE-205 02, Sweden; Intensive and Perioperative Care, Skåne University Hospital, Entrégatan 7, SE-222 42, Sweden; Anaesthesia and Intensive Care, Kristianstad Hospital, Kristianstad, SE-291 85, Sweden; Anaesthesia and Intensive Care, Helsingborg Hospital, Helsingborg, SE-251 87, Sweden; Department of Infectious Diseases, Skåne University Hospital, Malmö, SE-205 02, Sweden; Department of Translational Medicine, Lund University, Malmö, SE-221 00, Sweden; Neurology, Department of Clinical Sciences Lund, Skåne University Hospital, Lund, SE-221 00, Sweden

**Keywords:** COVID-19, health-related quality of life, post-intensive care syndrome, intensive care, post-acute COVID-19 syndrome

## Abstract

**Background:** Long-term outcomes after critical COVID-19 have not been sufficiently studied. This study aimed to describe changes in functional outcome and health-related quality of life (HRQoL) assessed at 3 and 12 months in a cohort of critically ill COVID-19 survivors. A secondary aim was to investigate factors associated with good functional outcome and HRQoL at 12 months.

**Methods:** This prospective multicentre cohort study included critically ill COVID-19 patients admitted to six intensive care units in Sweden between May 2020 and May 2021. Surviving patients were invited to face-to-face follow-ups at 3 and 12 months. A good functional outcome was a Glasgow outcome scale extended ≥7. HRQoL was assessed by the physical and mental component summary of the SF-36v2®, with T-scores ≥45 representing a good HRQoL. Factors associated with good functional outcome and HRQoL at 12 months were explored by multivariable logistic regression.

**Results:** A good functional outcome was found in 93/264 (35%) and 138/217 (64%) of survivors at 3 and 12 months, respectively. There was a significant improvement in the SF-36v2® Physical component summary (PCS) between 3 and 12 months (mean 40 versus 44, p*<*0.001). The SF-36v2® Mental component summary (MCS) was within the normal range at 3 months, with no significant change at 12 months (mean 46 versus 48, p=0.05). Older age was associated with a good functional outcome. Low clinical frailty and absence of diabetes mellitus were associated with a good physical HRQoL. A shorter duration of mechanical ventilation was associated with a good outcome for all three outcome measures.

**Conclusion:** Between 3 and 12 months, functional outcome and physical aspects of HRQoL significantly improved, indicating continued recovery up to at least one year after critical COVID-19. Low frailty, less comorbidity, and shorter duration of mechanical ventilation were associated with better long-term outcomes, while old age was associated with better functional outcome.

**Study registration:** ClinicalTrials.gov Identifier: NCT04974775, registered April 28, 2020.

## Introduction

Many patients with critical coronavirus disease 2019 (COVID-19) require intensive care [1, 2, 3], most of whom fulfil the Berlin definition of acute respiratory distress syndrome (ARDS) [4, 5]. Previous studies on intensive care survivors have described symptoms and sequelae known as post-intensive care syndrome (PICS) [6]. PICS refers to newly developed or worsening pre-existing symptoms in mental health, cognitive, and physical function and is associated with increased one-year mortality as well as a lower health-related quality of life (HRQoL) [7, 8]. Studies on survivors of ARDS from other causes than COVID-19 have shown a high prevalence of functional and cognitive impairments affecting the long-term HRQoL after intensive care [9, 10]. In a meta-analysis by Dowdy et al. [11], HRQoL remained persistently lower up to 4 years post-ARDS, especially in the physical domains, compared to a healthy population.

Irrespective of COVID-19 severity, an increasing number of studies report persisting symptoms and delayed long-term complications after acute infection, referred to as post-acute COVID-19 syndrome [12, 13]. Patients surviving critical COVID-19 may thus develop lasting impairment due to both PICS and post-acute COVID-19 [14, 15]. Cross-sectional telephone interviews at 4-6 months after the ICU stay show that critically ill COVID-19 survivors commonly report new disabilities [16]. However, COVID-19-related ICU stays were not associated with a lower quality of life in elderly patients compared with other causes of ICU stays [17, 18]. A study on ICU-treated COVID-19 patients transferred to a rehabilitation facility and assessed at 2 and 12 months showed persisting altered HRQoL at 12 months [19]. In an early single-centre study, fatigue and muscle weakness were common in 1-year COVID-19 survivors post-ICU, whereas HRQoL had almost returned to normal at 12 months [20]. Old age, clinical frailty, and invasive mechanical ventilation have been associated with increased mortality in critically ill COVID-19 patients. In survivors, female sex, severe acute illness, comorbidities, and mechanical ventilation were associated with lower HRQoL and poor functional outcome. The impact of socioeconomic status has shown conflicting results [21, 22], while the strain on the healthcare system at a specific time, the ICU burden, may have played a role [23]. Studies on long-term outcomes in critically ill COVID-19 survivors are thus continuously emerging. No large study has, however, systematically presented data from repeated long-term face-to-face follow-ups, and few have focused on identifying predictors of good outcome.

The primary objective of this study was to describe changes over time in functional outcome and HRQoL assessed at face-to-face follow-ups at 3 and 12 months in a large cohort of critically ill COVID-19 survivors. A secondary objective was to investigate factors associated with good functional outcome and HRQoL at 12 months.

## Method

### Study population

This prospective observational multicentre cohort study is a part of the SweCrit COVID-19 study, which included all critically ill adult patients (≥18 years old) with laboratory-confirmed SARS-CoV-2 infection at six intensive care units (ICU) in the Skåne Region, in southern Sweden, between May 11, 2020, and May 10, 2021[23]. Written informed consent was collected from all participants on admission, before discharge, or at a follow-up visit up to one year later. For deceased patients, consent was presumed. Patients were excluded if COVID-19 was not the primary cause of ICU admission. This manuscript was prepared per the STROBE guidelines for observational studies [24]. The Swedish Ethical Review Authority approved the SweCrit COVID-19 study (Dnr: 2020-01955, 2020-03483, 2021-00655).

### Design

At 3 and 12 months after ICU admission, surviving participants were invited to a follow-up, performed primarily face-to-face but sometimes replaced by a telephone interview. To increase interrater reliability, all outcome assessors participated in mandatory training and were given a written manual to conduct the follow-up in a structured order. We used certified interpreters when participants were deemed non-fluent in Swedish.

#### Patient characteristics

Information on patient characteristics was collected during the intensive care stay and at the 3- and 12-month face-to-face follow-ups, which has been described previously [23]. Since we lacked detailed information on socioeconomic status, we used data collected at the follow-up as a proxy, including living situation, marital status, employment status before COVID-19, level of education, and native language. At 3 and 12 months, we also collected information on the participants’ current employment status and overall Life satisfaction. Life satisfaction was assessed by a single question from the World Values Survey: “All things considered, how satisfied are you with your life as a whole these days?” and a Visual Analogue Scale (VAS) range 1-10, where 1 means “completely dissatisfied” and 10 “completely satisfied”. According to the Organisation for Economic Co-operation and Development (OECD) Better Life Index [25], Sweden’s average Life satisfaction VAS scale score is 7.3, whereas the OECD average is 6.7.

### Outcome measures

#### Functional outcome

Functional outcome was assessed at 3 and 12 months by the clinician-reported Glasgow outcome scale extended (GOSE), an ordinal scale ranging from 1 to 8, where 1 represents dead, and 8 illustrates full recovery, as presented in Supplementary Figure 1 [26]. A GOSE ≥7 was considered a good functional outcome for this study.

#### HRQoL

To investigate HRQoL, the patient-reported Short form health survey version 2 (SF-36v2^®^) was used at 3 and 12 months [27]. This instrument is recommended as a core outcome measure evaluating ARDS survivors after hospital discharge [28]. A Swedish version has been validated against the original version. The SF-36v2^®^ comprises 36 items summarised into eight health domains: Physical functioning, Role physical, Bodily pain, General health, Vitality, Social Functioning, Role-Emotional, and Mental health. These eight health domains are further aggregated into two overall component summary scores: the Physical component summary (PCS) and the Mental component summary (MCS). Scores for each domain can range from 0 (worst) to 100 (best). All eight domains can also be presented as standardised T-scores. A T-score of 50 represents the norm mean in a 2009 US sample, and 10 equals one standard deviation. At a group level, a T-score *<*47 indicates impaired health; at an individual level, a T-score *<*45 indicates impaired health in that domain. PCS and MCS are reported as T-scores and have been demonstrated to have good discriminative validity for identifying differences between clinically meaningful groups [29]. We further present Minimally important difference (MID) values from the SF-36v2^®^ manual [30]. MID is the smallest change perceived as relevant by the patient. The MIDs retrieved from the manual have been determined by an anchor-based approach, meaning the SF-36v2^®^ scores were linked to an external criterion, e.g., performance status or patient- or physician-reported health ratings.

The primary outcome measure was any change in functional outcome between 3 and 12 months assessed by the Glasgow Outcome Scale Extended (GOSE).

Secondary outcome measures included changes in HRQoL assessed by Short Form Health Survey version 2 (SF-36v2^®^), employment status and overall Life satisfaction.

### Statistics

The study enrolment was based on a population-based design predetermined to close after one year, and no power analysis was performed. Continuous variables are presented as the medians with 25th and 75th percentiles (Q1-Q3). Categorical variables are expressed as numbers and percentages. Standardised T-scores of SF-36v2^®^ are presented with mean values and 95% Confidence intervals (CI). For comparing two variables on an ordinal scale and continuous variables on an interval scale, the Sign test was used for dependent groups, and the Wilcoxon signed-rank test was used for independent groups. The McNemar test was used for differences in categorical and binary variables between two dependent groups. For the SF-36v2^®^ domains and the component summary scores (MCS and PCS), the absolute mean difference in T-scores between 3 and 12 months was measured to determine if this reached the MID, in addition to statistical significance [30]. To explore factors associated with a good functional outcome at 12 months (GOSE ≥7) and HRQoL (PCS ≥ 45 and MCS ≥ 45), we chose variables previously associated with 90-day survival after critical COVID-19 and, in addition, the collected proxies for socioeconomic status and ICU burden. The investigated variables were *Characteristics prior to the critical COVID-19* : age, sex, Body mass index (BMI), Clinical frailty scale and Charlson comorbidity index, complicated diabetes mellitus, hypertension, and smoking status. *The severity of acute disease*: symptomatic days before ICU, Simplified acute physiology score 3 (SAPS3), arterial oxygen partial pressure ratio to fractional inspired oxygen, P_a_O_2_/F_i_O_2_ (P/F) day 1, P_a_CO_2,_ day 1, Sequential organ failure assessment (SOFA)- admission score, invasive mechanical ventilation (IMV), the logarithm base 10 of the duration of IMV, ICU length of stay, hospital length of stay, tracheostomy and need for continuous renal replacement therapy (CRRT). *Socioeconomic status*: native Swedish speaker, single household, level of education*>*12 years, and employment before COVID-19. *ICU burden* : we used the number of ICU-treated COVID-19 patients in the region on the day of admission.

Variables with less than 10 events or more than 30% missing values were excluded from further analysis. The remaining variables had missing values at random. They were imputed by multiple imputations from the Statistical Package for the Social Sciences (SPSS®) version 27 using the AUTO method with five imputation sets. Univariable regression analyses were performed in the first step with GOSE ≥7, PCS ≥45 and MCS ≥45 at 12 months as dependent variables. In line with *purposeful selection* [31], we considered variables associated with a good functional outcome and HRQoL in the univariable analyses with a p-value *<*0.25 to enter the corresponding multivariable models. A correlation matrix investigated the selected variables to avoid multicollinearity. The Hosmer-Lemeshow test was used to evaluate the goodness of fit of the models. Multivariable logistic regressions were performed with the selected variables to explain a GOSE≥7, a PCS ≥45 and an MCS ≥45. We considered a p-value *<*0.05 statistically significant. Statistical analyses were performed using the SPSS^®^ version 27.

## Results

At 3 months, 303/498 patients were alive (61%), of whom 264/303 (87%) participated in the follow-up, and 175 (66%) were conducted face-to-face. At 12 months, 217/298 (73%) returned for a second follow-up, and 160 (74%) were conducted face-to-face.

A flow chart is presented in Figure 1. Demographics, admission characteristics, variables from intensive care and complications are summarised in Table 1, stratified by 3 and 12-month follow-up participants. Variables in survivors not participating in the follow-up at 12 months (86/303) were similar to the investigated participants aside from younger age (59 [46-66] vs 61 [52-68], p=0.03) and lower SAPS 3 (53 [42-65] vs 56 [47-65], p=0.01) (Supplementary Table 1). Baseline physical and psychological status and socioeconomic characteristics before COVID-19 were only available for survivors participating in the follow-ups.

**Fig. 1.**
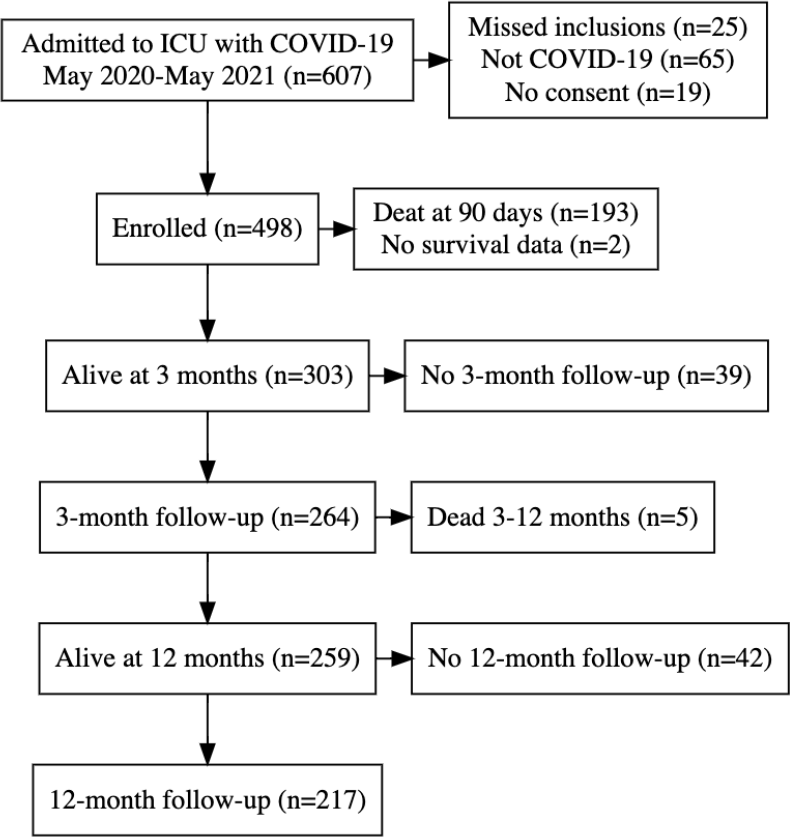
Flow chart at 12 months. Participants enrolled and completed 3- and 12-month follow-ups in this cohort of critically ill COVID-19 patients (n=498).

**Table 1.** Demographics and patient characteristics. Demographics, patient characteristics and socioeconomic status pre-COVID-19, ICU admission characteristics and complications from intensive care. Data was stratified on all patients, survivors at 90 days, and participants at 3- and 12-month follow-ups. Percentages for proportions. Median and (Q1-Q3) for continuous variables. ^a^Employment before ICU among participants at follow-up 20 to 64 years (n=106).^b^Minimum value of P/F (PaO_2_ / FiO_2_) in all patients (n=498) on day 1 in the ICU.^c^Maximum value of PaCO_2_ in all patients (n=498) on day 1 in the ICU

|  | Overall | Survivors 3 months | Patients 3 months | Patients 12 months | Complete data 12 months |
| --- | --- | --- | --- | --- | --- |
| *Demographics* |  |  |  |  |  |
| Number of patients | 498 | 303 | 264 | 217 | 217 |
| Age (years) | 66 [56-73] | 61 [52-68] | 61 [52-68] | 62 [53-69] | 100% |
| Male | 74% | 74% | 73% | 73% | 100% |
| *Patient characteristics pre-COVID-19* |  |  |  |  |  |
| BMI (kg/m<sup>2</sup>) | 30 [27-35] | 31 [27-36] | 31 [27-36] | 31 [27-35] | 100% |
| Smokers-ever | 44% | 38% | 37% | 38% | 100% |
| Charlson Comorbidity Index | 3 [2-4] | 2 [1-3] | 2 [1-3] | 2 [1-3] | 100% |
| Diabetes mellitus | 31% | 28% | 25% | 29% | 100% |
| Hypertension | 55% | 50% | 49% | 52% | 98% |
| COPD and severe asthma | 19% | 17% | 18% | 19% | 100% |
| Clinical Frailty Scale | 3 [2-4] | 3 [2-3] | 3 [2-3] | 3 [2-3] | 98% |
| *Socioeconomic characteristics pre-COVID-19 in follow-up cohort* |  |  |  |  |  |
| Native Swedish speaker | N/A | N/A | 58% | 60% | 100% |
| Single household | N/A | N/A | 24% | 24% | 99% |
| Level of education > 12 years | N/A | N/A | 36% | 36% | 98% |
| Employed before COVID-19<sup>a</sup> | N/A | N/A | 63% | 69% | 100% |
| *ICU admission characteristics and variables from intensive care and hospital stay* |  |  |  |  |  |
| Symptomatic days before ICU | 11 [8-15] | 10 [7-14] | 10 [7-14] | 11 [7-13] | 96% |
| ICU burden | 29 [18-34] | 27 [17-31] | 27 [17-31] | 28 [17-32] | 100% |
| SAPS3 | 60 [50-69] | 56 [47-65] | 57 [47-66] | 57 [48-66] | 100% |
| SOFA admission | 8 [5-9] | 7 [4-9] | 7 [4-9] | 8 [4-9] | 86% |
| P/F ratio Day 1 min<sup>b</sup> (kPa) | 9.0 [7.0-12] | 10 [7.0-13] | 10 [7.0-13] | 10 [7.0-13] | 90% |
| P<sub>a</sub>CO<sub>2</sub> Day 1 max<sup>c</sup> (kPa) | 5.8 [4.9-7.4] | 5.6 [4.8-6.7] | 5.7 [4.9-6.7] | 5.6 [4.9-6.7] | 91% |
| IMV | 72% | 66% | 67% | 68% | 100% |
| Duration of IMV (days) | 9.8 [5.2-19] | 8.3 [4.6-17] | 8.9 [4.5-18] | 9.0 [4.7-19] | 100% |
| ICU Length of stay (days) | 9.5 [4.9-17] | 9.5 [4.6-17] | 10 [5.0-19] | 9.2 [4.7-18] | 100% |
| Hospital length of stay (days) | 23 [15-42] | 23 [14-44] | 23 [14-45] | 23 [14-43] | 99% |
| Prone position | 80% | 80% | 80% | 75% | 100% |
| Tracheostomy | 15% | 13% | 12% | 12% | 100% |
| Sedation > 7 days | 93% | 88% | 88% | 87% | 100% |
| Neuromuscular blocking agents > 7 days | 19% | 12% | 13% | 13% | 54% |
| Vasopressors > 7 days | 47% | 39% | 39% | 38% | 65% |
| CRRT | 14% | 14% | 13% | 13% | 100% |
| ECMO | 3% | 2% | 3% | 3% | 100% |
| Pneumothorax | 11% | 7% | 7% | 6% | 80% |
| Pulmonary embolus | 17% | 15% | 15% | 15% | 100% |
| Resuscitation after cardiac arrest | 4% | 1% | 1% | 1% | 100% |

Among follow-up participants aged 20 to 64, 107/169 (63%) were employed before intensive care. At 3 months, 66/107 (62%) of those working before intensive care had returned to work, and the proportion at 12 months was 71/105 (68%). The mean Life satisfaction VAS scale score was 6.5 [6.2-6.9] (n=201) at 3 months and 7.0 [6.2-7.3] (n=199) at 12 months. At 12 months, 132/217 (61%) had received rehabilitation, and 34/217 (16%) had ongoing rehabilitation (Table 2).

**Table 2.** Functional outcome and HRQoL stratified by 3 and 12-month follow-up. The mean T-scores of the eight domains and the Physical component summary score (PCS) and Mental component summary score (MCS) of SF-36v2^®^. The difference in mean T-scores between 3 and 12 months, including reference values for Minimally Important Differences (MID) for each domain and summary score. Glasgow outcome scale extended (GOSE) and percentages returning to work, receiving rehabilitation and general Life satisfaction at 3 and 12 months. Sign test for ordinal and continuous variables. McNemar test for dependant categorical variables. Median and (Q1-Q3) for continuous variables. Mean and 95% CI in Life satisfaction and SF-36v2^®^variables. ^a^Minimally important difference. ^b^Comparison between the 12-month cohort (n=217) at 3 and 12 months. ^c^n=106 patients 20-64 years working before COVID-19. ^d^Mean Life satisfaction in Sweden was 7.3 in 2019.

| Outcome measure | 3-month follow-up (n=217) | 12-month follow-up (n=217) | Difference mean (MID<sup>a</sup>) | p-value<sup>b</sup> |
| --- | --- | --- | --- | --- |
| *T-scores of SF-36v2<sup>®</sup>* |  |  |  |  |
| PCS | 40 (39-42) | 44 (42-45) | 3.6 (2) | <0.001 |
| MCS | 46 (45-48) | 48 (47-50) | 1.9 (3) | 0.05 |
| Physical Functioning | 39 (38-41) | 44 (42-45) | 4.4 (3) | <0.001 |
| Role-Physical | 37 (35-38) | 43 (42-45) | 6.4 (3) | <0.001 |
| Bodily Pain | 44 (43-46) | 46 (44-48) | 1.6 (3) | 0.13 |
| General Health | 46 (45-48) | 46 (44-47) | -0.8 (2) | 0.42 |
| Vitality | 45 (43-47) | 47 (45-49) | 2.1 (2) | 0.05 |
| Social Functioning | 42 (40-44) | 47 (46-49) | 5.2 (3) | <0.001 |
| Role-Emotional | 42 (39-44) | 44 (42-46) | 2.7 (4) | 0.004 |
| Mental Health | 48 (46-49) | 49 (47-51) | 1.3 (3) | 0.10 |
| PCS ≥45 | 30% | 45% | N/A | 0.002 |
| MCS ≥45 | 56% | 65% | N/A | 0.11 |
| *Functional outcomes* |  |  |  |  |
| GOSE | 6 [5-7] | 7 [5-7] | N/A | <0.001 |
| Good recovery (GOSE ≥7) | 37% | 64% | N/A | <0.001 |
| Poor recovery (GOSE 3-4) | 20% | 7% | N/A | <0.001 |
| *Miscellaneous* |  |  |  |  |
| Return to work<sup>c</sup> | 52% | 68% | N/A | <0.001 |
| Rehabilitation post-COVID-19 | 54% | 61% | N/A | 0.006 |
| Ongoing rehabilitation post-COVID-19 | 32% | 16% | N/A | <0.001 |
| Life satisfaction (VAS scale)<sup>d</sup> | 6.5 (6.2-6.9) | 7.0 (6.2-7.3) | N/A | 0.003 |

### Functional outcome

At 3 and 12 months, 93/264 (35%) and 138/217 (64%) of patients had a good functional outcome (GOSE ≥7), respectively (Figure 2). The median GOSE was 6 [5-7] at 3 months, significantly increasing to 7 [6-7] at 12 months (p*<*0.001). Meanwhile, GOSE 3-4, indicating dependency in daily life, was found in 20% at 3 months, with a significant decrease to 7% at 12 months (p*<*0.001) (Table 2).

**Fig. 2.**
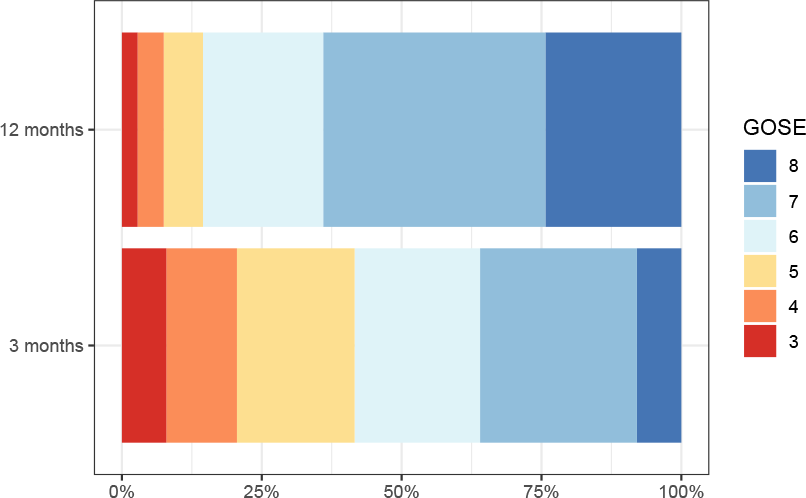
GOSE for survivors after critical COVID-19 at 3 and 12 months. Glasgow Outcome Scale Extended (GOSE) is an 8-grade ordinal scale measuring functional outcome, where 1 represents death and 8 full recovery. As only survivors are presented, and none scored GOSE 2 (vegetative state), percentages of participants’ GOSE are displayed as 3 to 8 at both 3 and 12 months.

### HRQoL

At 3 months, 221/264, and 12 months, 184/217, had completed the SF-36v2^®^. Participants not responding to the SF-36v2^®^ questionnaire at 12 months (119/264) were less often native Swedish speakers (43% versus 64%, p=0.002) compared to all follow-up participants but similar in other pre-disease characteristics (Supplementary Table 2).

The mean T-score of PCS was 40 (95% CI: 39-42) at 3 months, which increased to 44 (95% CI: 42-45) at 12 months (p*<*0.001). On an individual level, normal physical health (PCS ≥45) was observed in 30% of participants at 3 months and in 55% at 12 months (p=0.002). The mean T-score for MCS was 47 (95% CI: 45-48) at 3 months and 48 (95% CI: 47-50) at 12 months (p=0.05). On an individual level, normal mental health (MCS ≥ 45) was reported by 56% and 65% at 3 and 12 months, respectively (p=0.11). The change in the mean T-score of PCS (mean absolute difference: 3.6) between 3 and 12 months was above the threshold of MID (*>*2). The corresponding change in MCS (mean absolute difference: 1.9) was below the MID (*>*2) (Table 2 and Supplementary Table 3).

Of the eight SF-36v2^®^ domains, the lowest (worst) T-scores at 12 months were found in Physical functioning and Role physical. Conversely, the domains with the highest (best) T-scores at 12 months were Vitality, Social functioning, and Mental health, all within the normal range. The SF-36v2^®^ domains with a change between 3 and 12 months above a MID were Physical functioning, Role physical, and Social functioning (Table 2, Figure 3).

**Fig. 3.**
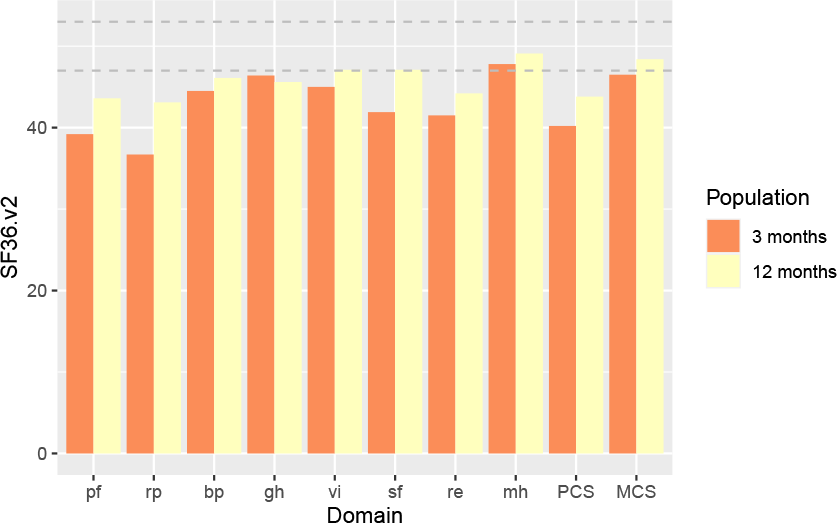
The SF-36v2^®^ mean T-scores of the 8 health domains and the Physical (PCS) and Mental (MCS) component summary at 3 and 12 months. The SF-36v2^®^ eight health domains are Physical functioning (PF), Role physical (RP), Bodily pain (BP), General health (GH), Vitality (VT), Social functioning (SF), Role emotional (RE), and Mental health (MH). The Physical component summary (PCS) and the Mental component summary (MCS) are aggregates of these 8 domains, presented as standardised T-scores. A T-score of 50 is the norm mean in a 2009 US sample. At a group level, a T-score *<*47 indicates impaired health, while a T-score *<*45 indicates poor health in that domain on an individual level.

### Variables associated with a good outcome

Variables associated with a good functional outcome and HRQoL at 12 months in univariable analyses are presented in Supplementary Table 4. In multivariable logistic regressions, the variables with significant association to good functional outcome (GOSE ≥7) at 12 months were increased age (OR 1.49, 95% CI 1.03-2.17.08, p=0.03 for every 10 years increase in age) and shorter duration of IMV (OR 0.24, 95% CI 0.08-0.71, p=0.01).

For HRQoL, the variables with a significant association to good physical health at 12 months (PCS ≥45) were: Clinical frailty scale (OR 0.52, 95% CI 0.29-0.90, p=0.02), absence of complicated diabetes mellitus (OR 0.10, 95% CI 0.01-0.89, p=0.04), and shorter duration of IMV (OR 0.20, 95% CI 0.06-0.66, p=0.008). Variables associated with good mental health at 12 months (MCS ≥45) were shorter duration of IMV (OR 0.32, 95% CI 0.11-0.96, p=0.04) and use of tracheostomy (OR 6.2, 95% CI 1.2-32, p=0.03) (Figure 4 and Supplementary Table 5).

**Fig. 4.**
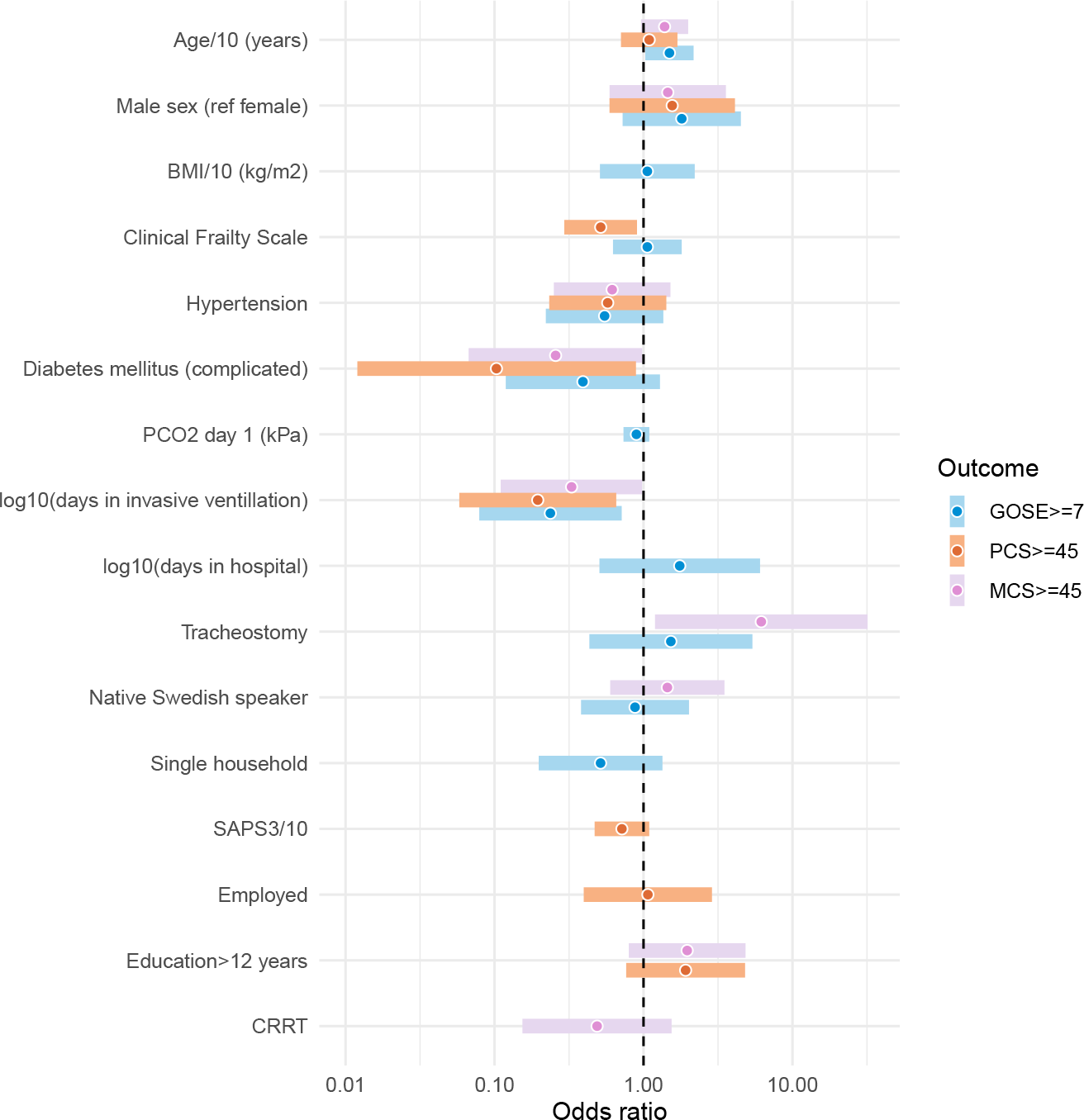
Forest plots of factors associated with good recovery at 12 months. Adjusted odds ratios with 95% CIs (demographics, comorbidities, acute physiology, ICU treatments, socioeconomic factors). The models were based on variables from the univariable logistic regression models. Variables with a p-value *<*0.25 were entered into the multivariable model. Light blue indicates Good functional outcome defined as Glasgow outcome scale extended (GOSE) ≥7 at 12 months. Light purple indicates Good physical HRQoL: T-score of the Physical component summary (PCS) ≥45 from SF-36v2^®^ at 12 months. Peach indicates Good mental HRQoL: T-score of the Mental component summary (MCS) ≥45 from SF-36v2^®^ at 12 months.

## Discussion

In this prospective multicentre cohort study, we found that survivors of critical COVID-19 significantly improved their functional outcome and physical aspects of HRQoL between 3 and 12 months. However, the mental aspects of HRQoL were already within the normal range at a group level at 3 months, with no significant further improvement at 12 months. In this face-to-face follow-up study, the results were consistent with previous studies in patients recovering from critical COVID-19, showing mild to moderate functional impairment early on and subsequent improvements within the first year [18, 32, 33]. In a recent study of critically ill COVID-19 patients conducted as an online survey, no significant recovery of PICS was seen between 12 and 24 months [34]. Our findings indicate, however, that recovery will continue for up to one year in critically ill COVID-19 survivors when face-to-face interviews are performed, which is a novel finding. Despite a general recovery, all the physical outcome domains of the SF-36v2^®^ remained below the average of a general US population at 12 months [30].

Risk factors of reduced HRQoL in survivors after non-COVID-19 ARDS include older age, the severity of disease, duration of IMV, neurocognitive dysfunction and comorbidities [35]. In follow-up studies on critically ill COVID-19 patients, the same factors were associated with a reduced HRQoL, underlining the similarities between ARDS in COVID-19 and ARDS from other aetiologies [18, 36, 37]. In the current study, we likewise found that a shorter duration of IMV was associated with good functional outcome and good physical and mental HRQoL at 12 months. Duration of IMV is a complex parameter that reflects the severity of the disease and the inherent impact of intensive care treatment, such as prolonged use of sedation and neuromuscular blocking agents. Interestingly, older patients had a better functional outcome, possibly due to decreased expectations in daily life. Furthermore, we found that the absence of diabetes mellitus and low clinical frailty were associated with good recovery of physical HRQoL. In addition, we found that tracheostomy was associated with a good mental HRQoL at 12 months. The latter finding might be incidental or reflect less sedative need in this group.

Our model did not associate young age and low frailty with a good mental HRQoL at 12 months. This may result from mental health being reported as relatively good at 3 months at a group level, and this model lacks granularity for this outcome. However, it may also reflect that information and variables on ICU admission are less effective in predicting mental HRQoL in the longer term and that other factors, such as access to rehabilitation or psychological support, maybe more important. Previous studies have shown conflicting results regarding the impact of socioeconomic status on outcome. A low socioeconomic status predicted poor clinical outcomes in one study but was associated with decreased hospital mortality in another study [21, 22]. In the present study, many participants did not have Swedish as their native tongue, and the level of education was well below the average in Sweden [38]. This could indicate an overall increased risk of ICU admission due to severe COVID-19 in these vulnerable groups [22]. However, we found no association between socioeconomic status and long-term outcomes in the adjusted model. Furthermore, we found no association between employment before COVID-19 and a good functional outcome. In previous studies, return to work corresponded well with functional recovery and could represent a proxy for good functional outcome.

Due to the general interest in post-COVID in society, novel rehabilitation pathways were created in many healthcare systems, including ours. Most of the participants in the present study had received rehabilitation during their first year, which may reflect that it was more readily available. Since we lacked detailed information about the interventions’ length, type, and intensity, no conclusion on the effect of the rehabilitation could be reached in our study. In general, participants who had received rehabilitation had worse functional outcomes at 3 and 12 months.

The strengths of this study include the prospective, multicentre design and the relatively large cohort of critically ill COVID-19 patients, who were studied for over 12 months. We assessed participants twice post-discharge, allowing temporal comparisons and analyses of the recovery trajectory. Conducting the follow-up interviews primarily face-to-face was another strength. Compared to telephone follow-ups used in most previous studies, in-person interviews made it possible to add missing data, which improved the overall quality [26]. As a result, we included most survivors at 3 and 12 months after ICU admission and with almost complete data sets, which allowed us to explore predictors of a good outcome. Face-to-face follow-ups also improve the sensitivity of the clinician-reported functional outcome [26]. Our group’s previous study on this cohort found that ICU burden was independently associated with 3-month mortality [23]. Including patients for one entire year thus allowed for exploring differences in functional outcomes in survivors due to strain on the intensive care organisation during the initial pandemic surges. We found no association between our definition of ICU burden and long-term recovery.

Some important limitations include the lack of baseline data on functional outcome and HRQoL due to the acute disease. It has been shown that HRQoL is substantially affected by impairments before the critical illness. As we lacked baseline data, we compared our results to a general US population used as a reference sample in the SF-36v2^®^ manual. This cohort, however, differs in age, sex, and disease burden and may not fully represent the baseline of our cohort. Still, we found the corresponding values from the Swedish SF-36v2^®^ population to be even less representative, underscoring the relevance of reporting patient characteristics associated with health status before ICU admission when studying long-term HRQoL. Furthermore, although using the MID references from the SF-36v2^®^ manual adds valuable information, they are primarily appropriate for groups with mean T-scores of 30-40. MIDs tend to be higher for the T-score ranges observed in this study, and the clinical implication must be interpreted with caution. Further, no adjustments for multiple testing were used due to the exploratory design. Also, this is a study on surviving patients after critical COVID-19, and with that comes a risk of survivorship bias.

## Conclusion

Two out of three survivors of critical COVID-19 had a good functional outcome assessed at a face-to-face interview 12 months after ICU admission. Impaired functional outcome and HRQoL were frequently reported at 3 months but improved significantly at 12 months. Low clinical frailty, fewer comorbidities, and shorter duration of IMV were independently associated with a good functional outcome and HRQoL at 12 months. Surprisingly, younger age was not associated with a good functional outcome or mental recovery in HRQoL.

## Data Availability

The datasets used and analysed during the current study
are available from the corresponding author upon reasonable
request

## List of abbreviations

ARDS: Acute respiratory distress syndrome
BMI: Body mass index
BP: Bodily pain
CI: Confidence interval
COVID-19: Coronavirus disease 2019
CRRT: Continuous renal replacement therapy
ECMO: Extracorporeal membrane oxygenation
GH: General Health
GOSE: Glasgow outcome scale-extended
HRQoL: Health-related quality of life
ICU: Intensive care unit
IMV: Invasive mechanical ventilation
LOS hospital: Hospital length of stay
LOS ICU: ICU length of stay
MID: Minimally important difference
MH: Mental health
MCS: Mental component summary
OECD: Organisation for Economic Co-operation and Development
OR: Odds ratio
P_a_CO_2_: Arterial partial pressure of carbon dioxide
P_a_O_2_: Arterial partial pressure of oxygen
P_a_O_2_/F_i_O_2_ (P/F): Arterial oxygen partial pressure ratio to fractional inspired oxygen
PCS: Physical component summary
PF: Physical functioning
PICS: Post-intensive care syndrome
RT-PCR: Real-time reverse transcriptase-polymerase chain reaction
RE: Role emotional
SAPS 3: Simplified acute physiology score 3
SARS-CoV-2: Severe acute respiratory syndrome Coronavirus 2
SF-36v2^®^: Short form health survey version 2
SIR: Swedish intensive care registry
SF: Social functioning
SOFA: Sequential organ failure assessment
SPSS: Statistical Package for the Social Sciences
STROBE: Strengthening the Reporting of Observational Studies in Epidemiology
VAS: Visual analogue scale
VT: Vitality
WHO: World Health Organization

## Declarations

### Ethical approval and consent to participate

Ethical approval was acquired from the Swedish Ethical Review Authority (2020/01955, 2020/03483 and 2020/05233). Written informed consent was obtained from all surviving participants on admission, before discharge or at the 90-day follow-up; for deceased patients, consent was presumed according to the approval from the Ethical review authority. The study was registered on ClinicalTrials.gov Identifier: NCT04974775, registered on April 28, 2020.

### Consent for publication

Not applicable

### Availability of data and materials

The datasets used and analysed during the current study are available from the corresponding author upon reasonable request.

### Competing interests

The authors declare that they have no competing interests.

### Funding

AF and HF were supported by governmental funding of clinical research within the Swedish National Health Service (ALF), regional funding from Region Skåne, the Swedish Heart-Lung Foundation and Skåne University Hospital Funds.

### Authors’ contributions

GL and HF designed the study. ID, ML, AL, and MS collected clinical data. GL coordinated and supervised the 3- and 12-month follow-ups. ID and AF performed the statistical analyses and prepared figures and tables. ID, GL, AF, and HF wrote the initial manuscript. All authors read and approved the final manuscript.

## Acknowledgements

We thank all patients and their next of kin who participated in this study and all staff at the ICUs of Skåne University Hospital in Malmö and Lund, Helsingborg Hospital and Kristianstad Hospital. Thank you to occupational therapists Anna Bjärnroos, Erik Mellerstedt, and Eva M Johnsson, physiotherapist Katarina Heimburg and research nurses Sara Andertun, Maria Nelderup, Anne Adolfsson, Marina Larsson, and Susann Schrey.

## Notes

### Competing Interest Statement

The authors have declared no competing interest.

### Author Declarations

The Swedish Ethical Review Authority approved the SweCrit COVID-19 study (Dnr: 2020-01955, 2020-03483, 2021-00655)

