## Supplemental tables and figures for "Factors influencing long-term recovery in critically ill COVID-19 survivors: A prospective multicentre cohort study"

**Table 1.** Participants missing from follow-up at 12 months compared to all survivors at 3 months. Medians and IQRs are shown for continuous variables. The Mann-Whitney U test was used for comparison between two independent groups. Percentages and Chi-square tests were used for proportions.

| Characteristic | 90-Day Survivors | Missing at 12-Month | p-value |
|---|---|---|---|
| No. of patients | 303 | 86 | N/A |
| Age (years) | 61 [52–68] | 59 [46–66] | 0.03 |
| Male sex | 74% | 76% | 0.62 |
| BMI (kg/m$^2$) | 31 [27–36] | 31 [27–36] | 0.93 |
| Charlson comorbidity index | 2 [1–3] | 2 [0.2–3] | 0.07 |
| Clinical frailty scale | 3 [2–3] | 3 [2–3] | 0.59 |
| Non-native Swedish speaker[a] | 64% | 48% | 0.12 |
| SOFA admission | 7 [4–9] | 7 [4–9] | 0.67 |
| SAPS3 | 56 [47–65] | 53 [42–65] | 0.01 |
| IMV | 66% | 63% | 0.46 |
| Duration of invasive ventilation (days) | 8.3 [4.6–17] | 7.0 [4.2–15] | 0.32 |
| ICU Length of stay (days) | 9.5 [4.6–17] | 9.5 [4.3–17] | 0.75 |

**Table 2.** Participants missing completing SF-36v2® at 12 months compared to all participants at the 3-month follow-up. Median and IQR for continuous variables. Mann-Whitney U for comparison between 2 independent groups. Percentage and Chi-square for proportions.

| Characteristic | 3-Month Follow-Up | Missing at 12 Months | p-value |
|---|---|---|---|
| No. of patients | 264 | 119 | N/A |
| Age (years) | 61 [52–68] | 59 [47–67] | 0.09 |
| Male sex | 73% | 73% | 0.99 |
| BMI (kg/m$^2$) | 31 [27–36] | 33 [28–36] | 0.27 |
| Charlson comorbidity index | 2 [1–3] | 2 [1–3] | 0.20 |
| Clinical frailty scale | 3 [2–3] | 3 [2–3] | 0.38 |
| Non-native Swedish speaker | 64% | 43% | 0.002 |
| SOFA admission | 7 [4–9] | 8 [4–10] | 0.58 |
| SAPS3 | 57 [47–66] | 54 [48–63] | 0.61 |
| IMV | 69% | 62% | 0.25 |
| Duration of IMV (days) | 8.9 [4.6–18] | 12 [7.3–26] | 0.07 |
| ICU length of stay (days) | 9.8 [5.0–19] | 8.3 [3.5–25] | 0.91 |

**Table 3.** Median 0-100 scores and the interquartile range of the eight SF-36v2® domains. Comparison between 3 and 12 months in SF-36v2® using the Mann-Whitney U test on the 184 available patients at 12 months. The U.S. norm refers to the 2009 norm and the Swedish norm to the 2004 norm.

| Domain | 3 Months | 12 Months | p-value | U.S. Norm | Swedish Norm |
|---|---|---|---|---|---|
| Physical Functioning | 55 [27–75] | 65 [40–85] | <0.001 | 80 | 88 |
| Role Physical | 44 [12–75] | 62 [41–88] | <0.001 | 80 | 86 |
| Bodily Pain | 51 [31–84] | 62 [41–100] | 0.13 | 70 | 73 |
| General Health | 58 [40–72] | 54 [39–77] | 0.42 | 65 | 74 |
| Vitality | 44 [31–62] | 50 [31–75] | 0.055 | 57 | 64 |
| Social Functioning | 62 [38–100] | 75 [50–100] | <0.001 | 82 | 86 |
| Role Emotional | 75 [33–100] | 83 [50–100] | 0.004 | 85 | 88 |
| Mental Health | 70 [50–85] | 75 [55–90] | 0.10 | 73 | 78 |

**Table 4.** Univariable regression for good functional outcome (GOSE $\geq$ 7), good physical HRQoL (PCS $\geq$ 45), and good mental HRQoL (MCS $\geq$ 45) at 12 months. Odds ratio (OR) and 95% Confidence interval (95% CI). Variables with a p-value <0.25 (grey row colour) were considered for the multivariable logistic regression analyses.

| Variable | p-value | OR | 95% CI Lower | 95% CI Upper |
|---|---|---|---|---|
| **GOSE $\geq$ 7 at 12 months** | | | | |
| Age/10 (years) | 0.24 | 1.15 | 0.92 | 1.43 |
| Male sex | 0.070 | 1.77 | 0.96 | 3.27 |
| BMI/10 (kg/m$^2$) | 0.065 | 0.65 | 0.41 | 1.03 |
| Ever smoker | 0.37 | 1.31 | 0.73 | 2.33 |
| Charlson comorbidity index | 0.86 | 1.02 | 0.84 | 1.24 |
| Clinical frailty scale | 0.21 | 0.81 | 0.58 | 1.13 |
| Diabetes mellitus (complicated) | 0.01 | 0.33 | 0.14 | 0.77 |
| Hypertension | 0.09 | 0.61 | 0.35 | 1.08 |
| Native Swedish speaker | 0.18 | 1.48 | 0.84 | 2.61 |
| Single household | 0.17 | 0.63 | 0.33 | 1.21 |
| Level of education >12 years | 0.26 | 1.41 | 0.77 | 2.59 |
| Employed before COVID-19 | 0.42 | 0.79 | 0.45 | 1.39 |
| Symptomatic days before ICU | 0.73 | 0.99 | 0.97 | 1.02 |
| SAPS3/10 | 0.31 | 0.89 | 0.70 | 1.12 |
| P/F ratio day 1 | 0.45 | 0.98 | 0.94 | 1.03 |
| PCO2p$CO_2$ day 1 | 0.17 | 0.90 | 0.77 | 1.05 |
| SOFA admission | 0.98 | 1.00 | 0.91 | 1.10 |
| IMV | 0.74 | 0.90 | 0.50 | 1.64 |
| log10(duration of IMV) (days) | 0.012 | 0.32 | 0.13 | 0.78 |
| Tracheostomy | 0.23 | 1.65 | 0.73 | 3.75 |
| CRRT | 0.59 | 0.81 | 0.39 | 1.71 |
| log10(hospital LOS) (days) | 0.22 | 1.62 | 0.75 | 3.46 |
| log10(ICU LOS) (days) | 0.75 | 0.91 | 0.50 | 1.65 |
| **PCS $\geq$ 45 at 12 months** | | | | |
| Age/10 (years) | 0.027 | 0.76 | 0.59 | 0.97 |
| Male sex | 0.22 | 1.50 | 0.77 | 3.00 |
| BMI/10 (kg/m$^2$) | 0.11 | 0.66 | 0.40 | 1.11 |
| Ever smoker | 0.36 | 0.75 | 0.41 | 1.40 |
| Charlson comorbidity index | 0.008 | 0.74 | 0.60 | 0.93 |
| Diabetes mellitus (complicated) | 0.009 | 0.12 | 0.02 | 0.58 |
| Hypertension | 0.002 | 0.39 | 0.21 | 0.71 |
| Clinical frailty scale | 0.010 | 0.58 | 0.38 | 0.87 |
| Native Swedish speaker | 0.74 | 1.10 | 0.58 | 2.10 |
| Single household | 0.54 | 1.20 | 0.62 | 2.00 |
| Level of education >12 years | 0.11 | 1.70 | 0.89 | 3.20 |
| Employed before COVID-19 | 0.095 | 1.70 | 0.91 | 3.00 |
| Symptomatic days before ICU | 0.39 | 0.98 | 0.95 | 1.00 |
| SAPS3/10 | 0.21 | 0.83 | 0.62 | 1.11 |
| P/F ratio day 1 | 0.61 | 1.00 | 0.96 | 1.10 |
| Pp$CO2_2$ day 1 | 0.89 | 0.99 | 0.83 | 1.20 |
| SOFA admission | 0.36 | 0.96 | 0.88 | 1.00 |
| IMV | 0.26 | 1.40 | 0.70 | 2.50 |
| log10(duration of IMV) (days) | 0.007 | 0.26 | 0.10 | 0.69 |
| Tracheostomy | 0.63 | 1.20 | 0.54 | 2.70 |
| CRRT | 0.43 | 0.74 | 0.35 | 1.60 |
| log10(hospital LOS) (days) | 0.90 | 0.95 | 0.42 | 2.16 |
| log10(ICU LOS) (days) | 0.26 | 0.67 | 0.35 | 1.26 |
| **MCS $\geq$ 45 at 12 months** | | | | |
| Age/10 (years) | 0.54 | 1.08 | 0.84 | 1.38 |
| Male sex | 0.25 | 0.74 | 0.44 | 1.24 |
| BMI/10 (kg/m$^2$) | 0.41 | 0.98 | 0.93 | 1.00 |
| Ever smoker | 0.83 | 1.10 | 0.53 | 2.20 |
| Charlson comorbidity index | 0.41 | 0.92 | 0.75 | 1.10 |
| Diabetes mellitus (complicated) | 0.006 | 0.27 | 0.11 | 0.68 |
| Hypertension | 0.23 | 0.69 | 0.38 | 1.30 |
| Clinical frailty scale | 0.379 | 0.84 | 0.57 | 1.20 |
| Native Swedish speaker | 0.059 | 1.80 | 0.98 | 3.20 |
| Single household | 0.93 | 1.00 | 0.52 | 2.00 |
| Level of education >12 years | 0.15 | 1.50 | 0.85 | 2.80 |
| Employed before COVID-19 | 0.61 | 1.20 | 0.64 | 2.10 |
| Symptomatic days before ICU | 0.51 | 1.00 | 0.96 | 1.10 |
| SAPS3/10 | 0.36 | 0.89 | 0.70 | 1.14 |
| P/F ratio day 1 | 0.5 | 0.98 | 0.93 | 1.00 |
| PCO2p$CO_2$ day 1 | 0.46 | 0.94 | 0.79 | 1.10 |
| SOFA admission | 0.84 | 0.99 | 0.90 | 1.10 |
| IMV | 0.93 | 1.00 | 0.55 | 1.90 |
| log10(duration of IMV) (days) | 0.041 | 0.38 | 0.15 | 0.96 |
| Tracheotomy | 0.086 | 2.30 | 0.89 | 5.90 |
| CRRT | 0.046 | 0.45 | 0.21 | 0.99 |
| log10(hospital LOS) (days) | 0.27 | 1.61 | 0.68 | 3.81 |
| log10(ICU LOS) (days) | 0.97 | 0.99 | 0.51 | 1.91 |

**Table 5.** Multivariable logistic regression for good recovery at 12 months. Good functional outcome: Glasgow outcome scale extended (GOSE) ≥7. Good physical HRQoL: T-score of Physical component summary from SF-36v2® (PCS) ≥45. Good mental HRQoL: T-score of Mental component summary from SF-36v2® (MCS) ≥45 Odds ratio (OR) and 95% Confidence intervals (95%CI).

| **Factor** | **OR [95% CI]** | **p-value** |
|---|---|---|
| **GOSE ≥7 at 12 Months** | | |
| Age /10 (years) | 1.5 [1.0–2.2] | 0.03 |
| Male sex | 1.8 [0.72–4.5] | 0.21 |
| BMI/10 (kg/m$^2$) | 1.1 [0.51–2.2] | 0.87 |
| Clinical frailty scale | 1.1 [0.62–1.8] | 0.83 |
| Hypertension | 0.55 [0.22–1.4] | 0.19 |
| Diabetes mellitus (complicated) | 0.39 [0.12–1.3] | 0.12 |
| PaCO2 day 1 (kPa) | 0.9 [0.73–1.1] | 0.28 |
| log10(duration of IMV) (days) | 0.24 [0.08–0.71] | 0.010 |
| log10(hospital length of stay) (days) | 1.8 [0.51–6.1] | 0.38 |
| Tracheostomy | 1.5 [0.43–5.4] | 0.51 |
| Native Swedish speaker | 0.88 [0.38–2.0] | 0.76 |
| Single household | 0.52 [0.20–1.3] | 0.17 |
| **PCS ≥45 at 12 Months** | | |
| Age/10 (years) | 1.1 [0.71–1.7] | 0.69 |
| Male sex | 1.6 [0.59–4.1] | 0.37 |
| Hypertension | 0.58 [0.23–1.4] | 0.23 |
| Diabetes mellitus (complicated) | 0.10 [0.01–0.89] | 0.040 |
| Clinical frailty scale | 0.52 [0.29–0.90] | 0.020 |
| SAPS3 points/10 | 0.72 [0.47–1.1] | 0.12 |
| log10 (duration of IMV) (days) | 0.20 [0.06–0.66] | 0.008 |
| Employed before COVID-19 | 1.1 [0.40–2.9] | 0.89 |
| Level of education >12 years | 1.9 [0.76–4.8] | 0.17 |
| **MCS ≥45 at 12 Months** | | |
| Age/10 (years) | 1.4 [0.95–2.0] | 0.088 |
| Male sex | 1.4 [0.59–3.5] | 0.43 |
| Hypertension | 0.57 [0.23–1.5] | 0.25 |
| Diabetes mellitus (complicated) | 0.28 [0.071–1.0] | 0.068 |
| log10(duration of IMV) (days) | 0.32 [0.11–0.96] | 0.041 |
| Tracheostomy | 6.2 [1.20–32] | 0.029 |
| CRRT | 0.46 [0.14–1.49] | 0.20 |
| Native Swedish speaker | 1.4 [0.60–3.5] | 0.42 |
| Level of education >12 years | 2.0 [0.79–4.8] | 0.15 |
| ICU burden/10 | 1.4 [0.84–1.4] | 0.58 |

. **Figure 1.** The 8-tiered Glasgow Outcome Scale Extended (GOSE)

| GOSE | Category | Description |
|---|---|---|
| 1 | Dead | |
| 2 | Vegetative State | Unawareness, only reflex responses, periods of eye-opening. |
| 3 | Lower Severe Disability | Entirely dependent for daily needs. Constant assistance required. Cannot be left alone. |
| 4 | Upper Severe Disability | Requires frequent care. Can be left alone for short periods. |
| 5 | Lower Moderate Disability | Can engage in simple activities. Limited social and leisure activity. |
| 6 | Upper Moderate Disability | Working reduced hours. Moderately active socially. |
| 7 | Lower Good Recovery | Capable of independent living. Work capability slightly reduced. |
| 8 | Upper Good Recovery | Functionally independent. Full participation in daily activities. |
